# Blood Immuno-metabolic Biomarker Signatures of Depression and Affective Symptoms in Young Adults

**DOI:** 10.1101/2024.06.03.24308351

**Authors:** Nicholas A Donnelly, Ruby SM Tsang, Éimear M Foley, Holly Fraser, Aimee L Hanson, Golam M Khandaker

## Abstract

**Background:** Depression is associated with alterations in immuno-metabolic biomarkers, but it remains unclear whether these alterations are limited to specific markers, and whether there are subtypes of depression and depressive symptoms which are associated with specific patterns of immuno-metabolic dysfunction.

**Methods:** To investigate whether immuno-metabolic biomarkers could be used to profile subtypes of depression, we applied regression, clustering, and machine learning to a dataset comprising depression diagnosis, depressive and anxiety symptoms, and blood-based immunological and metabolic biomarkers (n=118). We measured inflammatory proteins, cell count, lipids, hormones, and metabolites from up to n=4161 participants (2363 female, 337 with depression) aged 24 years from the Avon Longitudinal Study of Parents and Children birth cohort.

**Results:** Depression at age 24 was associated with both altered concentrations of immuno-metabolic markers, and increased extreme-valued inflammatory markers. Inflammatory and metabolic biomarkers show distinct, opposing associations with somatic and anxiety symptoms. We identified two latent components representing the relationship between blood biomarkers, symptoms, and covariates, one characterised by higher somatic symptoms and inflammatory markers (neutrophils, WBC, IL-6), and the other characterised by higher anxiety and worry and lower inflammatory markers (CRP, WBC, IL-6). Individuals with higher somatic-inflammatory component scores had greater depressive symptoms severity over the next five years. Immuno-metabolic biomarkers predicted depression diagnosis (Balanced Accuracy=0.580) and depression with high somatic symptoms (Balanced Accuracy=0.575) better than chance, but not depression with high anxiety symptoms (Balanced Accuracy=0.479).

**Conclusions:** Alterations in immuno-metabolic homeostasis is present in young adults with depression well before the typical age of onset of cardiometabolic diseases. The relationships between affective symptoms and blood immunometabolic biomarkers indicate two biotypes of depressive symptoms (somatic-inflamed vs anxious-non-inflamed). These patterns are relevant for prognosis and prediction, highlighting the potential usefulness of immuno-metabolic biomarkers for depression subtyping.

**Highlights:**

- Immuno-metabolic biomarkers show varying, opposing associations with somatic, mood and anxiety symptoms.
- Depression diagnosis and fatigue are associated with increased extreme-valued inflammatory markers.
- Two latent components (somatic-inflamed *vs* anxious-non-inflamed) represent the relationship between biomarkers, symptoms and key covariates.
- Higher somatic-inflamed scores are associated with greater depressive symptom severity in follow-up.
- Immuno-metabolic biomarkers predict depression and depression with high somatic symptoms, but not high anxiety symptoms, better than chance.

## 1. Introduction

The presence of immunological and metabolic alterations in depression has been widely replicated, including changes in the concentrations of blood inflammatory biomarkers (e.g., cytokines, acute phase proteins and immune cell counts (Foley et al., 2023; Frank et al., 2021; Goldsmith et al., 2016; Lynall et al., 2020; Osimo et al., 2020)) and metabolic biomarkers (e.g., lipids and metabolites (Bot et al., 2020; Davyson et al., 2023)).

Existing case-control studies have typically focused on specific biomarker domains (e.g., proteins, metabolic biomarkers, cells). Studies involving broader sets of biomarkers could clarify if immuno-metabolic alterations in depression are restricted to a few biomarkers or reflect broader disruption of immuno-metabolic homeostasis. Studies also typically compare mean biomarker levels between case and control groups, and often overlook substantial individual variability in biomarkers, which can provide important additional information regarding the overall pattern of change (Osimo et al., 2020). Recent neuroimaging findings suggest that depression and other psychiatric disorders are characterised by increased total number of extreme deviations from normative models of grey matter volume (Segal et al., 2023), but it is unclear whether such extreme variability is present in immuno-metabolic measures.

Depression is a syndrome comprising core symptoms of low mood and anhedonia, alongside depressive cognitions (e.g., guilt and hopelessness), anxiety symptoms, and somatic symptoms (e.g. fatigue, and changes to sleep and appetite). It is possible that immuno-metabolic processes have differing relationships to different symptoms (Milaneschi et al., 2021). Published symptom-level investigations have involved few specific inflammatory markers, most often interleukin 6 (IL-6) and C-reactive protein (CRP). These biomarkers appear to be particularly associated with somatic, atypical or energy-related symptom domains which include increased sleep, increased weight, fatigue, anhedonia and reduced motivation (Chu et al., 2019; Frank et al., 2021; Milaneschi et al., 2021), indicating that particular depression symptom profiles could be associated with specific immuno-metabolic biomarker patterns.

By examining the association between a broad set of immuno-metabolic biomarkers and the spectrum of depressive presentations, from individual symptoms to symptom domains and overall diagnosis, it may be possible to identify distinct clusters of biomarker-symptoms associations. Such associations could infer biomarker-based depression biotypes that might inform prognosis and treatment decisions, similar to those proposed using brain imaging data (Tozzi et al., 2024).

We report an in-depth exploration of the relationship of blood immuno-metabolic biomarkers (118 inflammatory proteins, cell count, lipids, and hormones) and depression measures (ICD-10 depression diagnosis, 4 symptom domains and 22 symptoms) based on data from young adults aged 24 years from the Avon Longitudinal Study of Parents and Children (Northstone et al., 2019), a UK birth cohort. In particular, we aimed to: (1) examine the pattern of immuno-metabolic biomarker associated with ICD-10 depression compared to controls, including extreme values of biomarkers; (2) quantify relationships between individual symptoms and immuno-metabolic variables, and explore if these associations formed distinct clusters or latent components; and (3) investigate whether machine learning models trained on immuno-metabolic variables can predict ICD-10 diagnoses of depression and specific symptom profiles within depression identified using the symptom clustering approach, comparing model performance with models trained on sociodemographic and clinical data.

## 2. Materials and Methods

### 2.1 Datasets

#### 2.1.1 Cohort Profile

We used data from the Avon Longitudinal Study of Parents and Children (ALSPAC) birth cohort (Boyd et al., 2013; Fraser et al., 2013; Northstone et al., 2019). This cohort study recruited pregnant women in the Avon region of England between 1990 and 1992 (Golding et al., 2001). After all rounds of recruitment, the final number of people recruited as children in study who were alive at age 1 was 14,901.

Participants in the study have been followed up with regular clinical and questionnaire-based assessments, and over 80,000 variables are available. Study data are captured and managed using the REDCap tool, hosted at the University of Bristol (Harris et al., 2019, 2009). Variables can be explored through a fully searchable data dictionary (http://www.bristol.ac.uk/alspac/researchers/our-data/), and data is available on a managed open access basis.

Ethical approval for the study was obtained from the ALSPAC Ethics and Law Committee and Local Research Ethics Committees. All participants provided written informed consent. Biological samples were collected with informed consent and in accordance with the UK Human Tissue Act (2004).

#### 2.1.2 Mental Health Data

Our main outcomes for our cross-sectional analysis were ICD-10 depression diagnosis and specific symptoms and symptom domains derived from the Clinical Interview Schedule – Revised (CIS-R (Lewis et al., 1992)) and the Community Assessment of Psychic Experiences (CAPE (Mossaheb et al., 2012)) questionnaires at age 24.

Given previous research suggesting that depressive symptoms can be best described by a “psychological” and a “somatic” factor (Thorp et al., 2020), we derived a domain score for psychological (0–4 scale) and somatic (0-9 scale) symptoms. For comparison to previous studies (Donnelly et al., 2022; Lamers et al., 2020; Milaneschi et al., 2021) we also created atypical (0–5 scale) and anxiety (0–7 scale) symptom domain scores.

We derived individual depression symptoms, combining items from the CIS-R and CAPE to target a wide set of symptoms, including those previously been associated with inflammation and metabolic dysfunction. This produced a total of 22 individual items: Aches & Pains (ach), Anhedonia (anh), Anxiety (anx), Appetite Decreased (app_dec), Appetite Increased (app_inc), Concentration Impaired (con), Depressive Cognitions – feelings of guilt, inadequacy and helplessness (dcog), Decreased interested in sex (dsx), Fatigue (ftg), Irritability (irt), Morning low mood (morn), Motivation impairment (mot), Panic (pan), Phobias/specific fears (pho), Restlessness (rstl), Sadness/low mood (sad), Slowed movements (slow), Sleep Decreased (slp_dec), Sleep Increased (slp_inc), Self-neglect (sneg), Suicidal Thoughts (stb), and Worry (wor). Full details of score derivations including ALSPAC variable codes used are given in the **Supplementary Methods**.

We used the Short Mood and Feeling Questionnaire (SMFQ (Rhew et al., 2010)) total score, completed by ALSPAC participants at four timepoints between ages 25 and 30, as outcomes in prospective longitudinal analysis.

#### 2.1.3 Blood Biomarkers

As exposures we included a set of immune proteins measured by the Olink Target 96 Inflammation Panel (Goulding et al., 2022) alongside blood cell counts, lipids, hormones and metabolites measured at age 24 (**Supplementary Table 1**). Blood samples were taken during F24 clinic visits in lithium heparin tubes, which were placed on ice immediately, spun down, plasma extracted and frozen at −80°C within 90 minutes. A total of 3027 blood samples from the F24 clinic were processed by Olink. The Olink Target 96 Inflammation panel detects a set 92 immune response-related proteins, returning Olink Proteomics’ arbitrary normalised protein expression (NPX) values on a log_2_ scale. Within the ALSPAC sample, a correlation of 0.77 has been demonstrated between a clinical chemistry assay and the Olink measure of Interleukin-6, indicating consistency between the measures (Goulding et al., 2022). We combined the Olink data with blood test results from the original F24 clinic, giving a total of 118 immuno-metabolic variables, based on having >90% complete data.

We included all Olink measures regardless of limit of detection as all measurements may have biological relevance. For multiple imputation analysis we used Olink data from 3005 individuals who had the same variables measured at age 9 (of whom 1826 had measurements at both age 9 and 24, see **Supplementary Methods)**.

### 2.2 Statistical Analysis

#### 2.2.1 Software

Statistical Analysis was carried out using R Statistical Software (version 4.4.2 (R Development Core Team, 2017)).

### 22.2 Immuno-metabolic Variable Selection and Preparation

All Olink data were expressed as Normalised Protein Expression (NPX values) on a log_2_ scale, therefore, no further transformations were performed. Non-Olink blood variables were log_2_ transformed prior to analysis. All immuno-metabolic variables were z-transformed by subtracting the overall mean value and dividing by the standard deviation, such that a 1-unit change in model results represents a 1 standard deviation (SD) unit change in the immuno-metabolic variable. Z-transformation was used to allow comparison between all immuno-metabolic variables on the same scale, and for consistency of model fitting.

#### 2.2.3 Univariate Statistical Models

We identified potential covariates using a directed acyclic graph ((Tennant et al., 2021), **Supplementary Methods**): sex assigned at birth (reference group=male), BMI at age 24, daily smoking (reference group=non-smoker), alcohol use (measured via the AUDIT-C questionnaire) and self-reported physical health problems (combining reported diabetes, asthma or COPD, arthritis, heart disease or heart problems, stroke, cancer or kidney disease into a single binary variable indicating absence or presence). In line with ALSPAC guidance when tabulating covariates, cell counts of less than 5 are replaced with “<5”.

For each immuno-metabolic variable, we fit two statistical models: (1) unadjusted model: depression diagnosis as outcome and the immuno-metabolic variable as predictor, (2) an adjusted model adding all covariates. Models were fit using the *glm* function, and average marginal effects (AME) calculated using the *marginaleffects* package, where we calculated the average change in probability of depression with a 1 SD unit increase in the immuno-metabolic variable. We were interested in both positive and negative changes in risk of depression. Given our large number of immuno-metabolic variables, we controlled the false discovery rate using the Bioconductor *swfdr* package (Boca and Leek, 2018; Korthauer et al., 2019). As sensitivity analyses we investigated the impact of sex on the association between haemoglobin and depression by repeating our models stratified by sex at birth.

To investigate the impact of attrition from the cohort we used multiple imputation (MI) to impute missing data in our age 24 dataset, using the R *mice* package and the Random Forest method. MI was performed on data from participants who either had blood sample data processed by Olink at age 9 or age 24 (total n = 4164). All age 24 blood variables and CIS-R-derived outcomes were imputed. For full details, see **Supplementary Methods**.

#### 2.2.3 Symptom-Biomarker Association

We fit models predicting depression measures (diagnosis, domain scores and symptom scores) using immuno-metabolic variables and the full set of covariates (above) for all pairs of depression- and immuno-metabolic measures. Models were fit with a binomial distribution using *glm*. We then took the matrix of AME values for all pairs of symptoms and immuno-metabolic variables and applied the *hclust* function in the R stats package, using Euclidean distance and the “complete” method. The resulting dendrogram indicated that maximum separation between variables was achieved by a 3-cluster solution. Clusters were manually labelled after inspecting their constituent associations.

#### 2.2.5 Extreme Value Analysis

We explored whether a given immuno-metabolic variable was extreme valued by taking non-depressed participants and calculating a normative model for each variable (using *lm* and our full set of covariates as predictors). Then, for each participant (both non-depressed and depressed) a standardised residual was calculated using the model fit to non-depressed individuals. A standardised residual was classified as “extreme” if the absolute value was greater than 2.6, based on a study applying a similar approach to brain imaging (Segal et al., 2023). We then took the sum of extreme values across immuno-metabolic variables for each individual.

Models predicting the count of extreme values from depression diagnosis or depression symptoms were fit with all covariates using a Poisson mixed effects model with participant ID as a random intercept.

#### 2.2.6 Partial Least Squares

We fit principal components analysis (PCA) and partial least squares models (PLS) to depressive symptom scores and biomarker variables (from the positive and negative association clusters) using the mixOmics package. PLS was performed using the *spls* function (Rohart et al., 2017) in “regression” mode. All covariates were included in PLS models. We selected 2 latent components for further analysis after applying a cross-validation approach, inspecting model performance with 1–5 components. A 2-component solution was optimal based on inspecting the Q_2_ statistic. We next used cross-validation to select the number of variables for each component, selecting variables using the “cor” measure.

We extracted PLS component scores for all components and individuals and used the scores as predictors of the total sum score on subsequent SMFQ measurements aged 25 - 30, using a generalised linear mixed model with a poisson distribution, fit with the *glmm* function in the lme4 package.

#### 2.2.7 Machine Learning Model Fitting

We fit elastic net regression models with the *glmnet* R package to predict ICD-10 depression diagnosis. In addition, we assessed predictive performance of two symptom profiles within participants with ICD-10 depression: depression *with* high levels of anxiety (high anxiety and worry, sum score ≥ 5) or depression *with* high somatic symptoms (fatigue, decreased sleep, low motivation, and aches & pains; sum score ≥ 9). The cut-offs were chosen to produce balanced groups based on the distributions of symptom profile scores.

We used four sets of predictors: (1) *Immuno-metabolic*: positive and negative-association cluster variables, plus extreme value counts and covariates; and three sets of comparison predictors: (2) *Sociodemographic*: all covariates, plus maternal occupational social class and history of adverse childhood events aged 0–17 (Houtepen et al., 2018); (3) *Mental Health History*: previous CIS-R depression diagnosis at age 18, SMFQ total score at ages 10 to 23, Strengths and Difficulties (SDQ) Total Difficulties Score at ages 7 and 9, and Maternal Edinburgh Postnatal Depression Scale Score; (4) *Full*: all variables included in previous models. Nested cross-validation (20 outer folds, 20 inner folds) was used to tune model hyperparameters (penalty and mixture parameters, see **Supplementary Methods**).

Model performance was measured using balanced accuracy (Sensitivity + Specificity) / 2 to allow comparison to similar analyses (e.g. (Winter et al., 2024)), evaluated by fitting a model to the held-out test data on each outer fold, for all 20 outer loops and calculating the mean and 95% confidence interval of these performance results.

## 3. Results

### 3.1 Cohort Profile

The complete case dataset contained 2954 individuals (Male N=1082, Female N=1872), of whom 309 (10.5%) had ICD-10 depression at age 24 (**Table 1** and **Supplementary Figure 1B-G**).

**Table 1:** Characteristics of ALSPAC study sample.

| Variable | Overall<br>N = 2,954<sup>1</sup> | Sex at Birth |  |
| --- | --- | --- | --- |
|  |  | Male | Female |
|  |  | N = 1,082<sup>1</sup> | N = 1,872<sup>1</sup> |
| ICD-10 Depression |  |  |  |
| No Depression | 2,645 (90%) | 1,011 (93%) | 1,634 (87%) |
| Depression | 309 (10%) | 71 (6.6%) | 238 (13%) |
| BMI @ 24 | 23.6 (21.4, 26.8) | 24.1 [21.8, 26.8] | 23.3 [21.3, 26.8] |
| Missing | 28 | 6 | 22 |
| Daily Smoking @ 24 | 341 (12%) | 130 (12%) | 211 (11%) |
| Missing | 18 | 7 | 11 |
| AUDIT Alcohol Questionnaire Score @ 24 | 5.00 (4.00, 7.00) | 6.00 [4.00, 8.00] | 5.00 [3.00, 7.00] |
| Missing | 32 | 13 | 19 |
| **Self-reported physical health conditions** | 379 (13%) | 148 (14%) | 231 (13%) |
| **Missing** | 85 | 30 | 55 |
| **Self-reported health by type** |  |  |  |
| **Arthritis** | 16 (0.6%) | 5 (0.5%) | 11 (0.6%) |
| **Asthma or COPD** | 320 (11%) | 125 (12%) | 195 (11%) |
| **Diabetes** | (0.5%) | (0.9%) | - |
| **Heart Disease or Heart problems** | 23 (0.8%) | 6 (0.6%) | 17 (0.9%) |
| **Kidney Disease** | <5 | <5 | <5 |
| **Stroke or cancer** | <5 | <5 | <5 |
| **Missing** | 85 | 30 | 55 |
| <sup>1</sup> n (%); Median [Q1, Q3] note cell counts with n < 5 are replaced with "<5" or not shown in line with ALSPAC guidance. |  |  |  |

### 3.2 Biomarker-Depression Associations

We examined the associated between immuno-metabolic variable levels (n=118 immuno-metabolic variables) and risk of depression. The results represent the change in probability of depression associated with a 1 standard deviation unit change in the immuno-metabolic variable of interest, where a positive value indicates an increase in risk, and a negative value indicates a decrease in risk.

In unadjusted analysis, after correction for multiple comparisons, 11 blood measures were associated with depression (**Table 2**): Haematocrit (average marginal effect [95% confidence interval]) = −0.022 [−0.033, −0.011], Haemoglobin = −0.019 [−0.03, −0.008], Aspartate aminotransferase (AST) = −0.018 [−0.031, −0.005], Red blood cell count = −0.018 [−0.029, −0.007], Alanine aminotransferase (ALT) = −0.017 [−0.029, −0.005]), Platelets = 0.016 [0.006, 0.027], Insulin = 0.018 [0.007, 0.029], CUB domain-containing protein 1 (CDCP1) = 0.019 [0.009, 0.029], White blood cell (WBC) count = 0.021 [0.011, 0.031], Interleukin-6 (IL-6) = 0.021 [0.012, 0.031] and Neutrophil count = 0.022 [0.012, 0.032].

**Table 2:** Immuno-metabolic biomarker associations with ICD-10 depression at age 24 years in the ALSPAC cohort Footnote: Biomarkers were selected based on FDR-corrected P-Value (P_FDR_) of <0.05 in the unadjusted model. Values displayed are Average Marginal Effects (95% Confidence Interval); positive values indicate higher absolute risk of depression associated with a 1 standard deviation unit increase in that immuno-metabolic variable; negative values indicate a lower absolute risk of depression associated with a 1 standard deviation unit increase. See Supplementary Table 1 for results for all biomarkers.

| Immuno-metabolic Variable | Unadjusted | Adjusted |
| --- | --- | --- |
| **Alanine aminotransferase (ALT)** | -0.017 [-0.029, -0.005] | -0.015 [-0.028, -0.002] |
| **Aspartate aminotransferase (AST)** | -0.018 [-0.031, -0.005] | -0.01 [-0.024, 0.003] |
| **CUB domain-containing protein 1 (CDCP1)** | 0.019 [0.009, 0.029] | 0.011 [-0.001, 0.022] |
| **Haemoglobin (Hb)** | -0.019 [-0.03, -0.008] | -0.003 [-0.018, 0.013] |
| **Haematocrit (Hct)** | -0.022 [-0.033, -0.011] | -0.011 [-0.025, 0.003] |
| **Interleukin-6 (IL-6)** | 0.021 [0.012, 0.031] | 0.016 [0.004, 0.027] |
| **Insulin** | 0.018 [0.007, 0.029] | 0.007 [-0.006, 0.02] |
| **Neutrophils** | 0.022 [0.012, 0.032] | 0.013 [0.002, 0.024] |
| **Platelets** | 0.016 [0.006, 0.027] | 0.009 [-0.003, 0.021] |
| **Red blood cell count (RBC)** | -0.018 [-0.029, -0.007] | -0.006 [-0.022, 0.01] |
| White blood cell count (WBC) | 0.021 [0.011, 0.031] | 0.01 [-0.001, 0.021] |

Evidence for association remained for ALT, IL-6, and neutrophil count after adjustment for covariates (sex at birth, smoking, alcohol use, BMI and physical health conditions), though the respective P-Values did not survive correction for multiple testing. See **Supplementary Table 1** for results for all biomarkers.

We carried out a series of sensitivity analyses. To investigate the impact of missing data due to cohort attrition we used multiple imputation for missing data and fit the same models to the imputed data. The results showed similar patterns of association (**Supplementary Figure 2)**.

The observed association between haemoglobin and depression was explained by sex at birth. On average males had haemoglobin values 17.6 g/L higher than females, while females had a 5.87% higher rate of depression than males. There was no association between depression and haemoglobin when modelling stratified by sex; for males the average marginal effect of a 1 SD unit change in Hb was −0.001 [−0.017, 0.015] and for females −0.002 [−0.017, 0.014].

We adjusted our univariate models for self-reported physical health conditions. As some conditions were rare (e.g. diabetes), we were unable to test associations between individual conditions and depression. However, the overall prevalence of self-reported physical health conditions was not higher in participants with depression (12.1%) than in participants without (13.3%), Chi-Squared test Χ_df=1_= 0.268, P-Value = 0.604.

### 3.3 Symptom-Biomarker Association Clusters

We carried out exploratory analyses to investigate whether immuno-metabolic variables were associated with individual depression and anxiety symptoms (n=22) and symptom domains (n=4, **Supplementary Methods and Supplementary Figure 3)**, and whether immuno-metabolic variables could be clustered on these associations.

We found differing patterns of associations between immuno-metabolic variables and symptoms (**Figure 1**), with some immuno-metabolic biomarkers appearing to have positive associations with most symptoms (e.g. CCL25), whereas others appeared to have largely negative associations (e.g. ALT). Somatic or atypical symptoms or symptom domains as well as psychological symptoms had positive associations with many immuno-metabolic variables (i.e. higher levels of biomarkers were associated with *higher* probability of endorsing a symptom), whereas in general anxiety and worry had negative associations (i.e. higher levels of biomarkers were associated with *lower* probability of endorsing a symptom).

**Figure 1:**
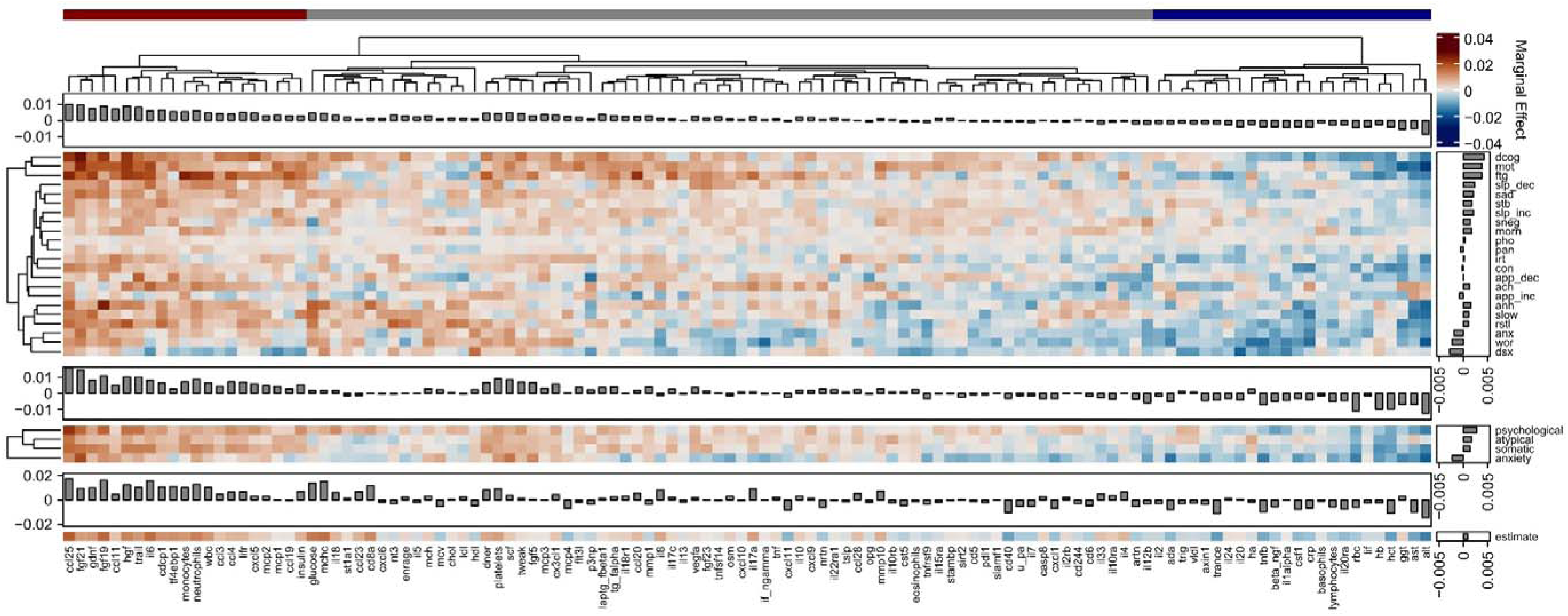
Associations of immuno-metabolic variables with depression and anxiety-related measures in the ALSPAC cohort Matrix plots showing the average marginal effect of a 1 SD-unit change in an immuno-metabolic variable on the probability of endorsing a symptom. From top to bottom, plots are of 22 individual depressive/anxiety symptom scores: Aches & Pains (ach), Anhedonia (anh), Anxiety (anx), Appetite Decreased (app_dec), Appetite Increased (app_inc), Concentration Impaired (con), Depressive Cognitions – feelings of guilt, inadequacy and helplessness (dcog), Decreased interested in sex (dsx), Fatigue (ftg), Irritability (irt), Morning low mood (morn), Motivation impairment (mot), Panic (pan), Phobias/specific fears (pho), Restlessness (rstl), Sadness/low mood (sad), Slowed movements (slow), Sleep Decreased (slp_dec), Sleep Increased (slp_inc), Self-neglect (sneg), Suicidal Thoughts (stb) and Worry (wor) (see **Supplementary Methods** for derivations); four depressive/anxiety symptom domain scores (anxiety, atypical, psychological and somatic) and ICD-10 depression diagnosis. Within each plot, marginal histograms represent the averaged marginal effect over all rows for each column in the plot (i.e., average over all outcomes to give an overall effect for each immuno-metabolic variable), marginal histograms to the right represent the averaged AME over all columns for each row in the plot (i.e. average over all immuno-metabolic variables for each outcome). At the top, the dendrogram from hierarchical clustering of the CIS-R symptom score matrix; immuno-metabolic variables are grouped based clustering of all mental health measures. Symptoms are also sorted by a similar clustering process. Note that all matrix plots are on the same colour scale. Coloured bars at the top indicate the cluster identity assigned to each immuno-metabolic variable: red = positive association; grey = low association; blue = negative association.

Hierarchical clustering applied to all pairwise associations of symptoms and immuno-metabolic variables identified three clusters, which were distinct with regards to their relationship to symptoms (see **Supplementary Table 1** for the cluster identity of all immuno-metabolic variables). Cluster 1 (*negative association*) comprised 24 immuno-metabolic variables, including some immune cell counts (lymphocytes, basophils), liver enzymes (ALT, AST and γGT), metabolic markers (Triglycerides, VLDL) and markers of red blood cell function (Hct, Hb, RBC), alongside cytokines that showed negative associations with symptoms, particularly anxiety and worry. Cluster 3 (*positive association*) comprised 21 biomarkers which included cytokines (e.g., IL6), chemokines (e.g., CCL4 and CCL25) immune cell counts (e.g., neutrophils, monocytes and white blood cells) and growth factors (FGF19, FGF21, HGF, GDNF). Variables in this cluster showed positive associations with specific somatic and psychological symptoms (including depressive cognitions, impaired motivation, fatigue, and decreased sleep). Cluster 2 (*low association*) comprised 73 biomarkers with predominantly small associations with symptoms.

### 3.4 Extreme Valued Biomarkers in Depression

We carried out an exploratory analysis to test if depression diagnosis was associated with increases in the variability of immuno-metabolic markers, which could be indicative of systemic immune or metabolic dysfunction. We calculated the total number of extreme-valued measurements for each immuno-metabolic variable and each participant.

All biomarkers had at least one participant with an extreme value; positive-signed extreme values were more common than negative-signed values (Figure 2A). After adjusting for covariates, participants with depression had higher total extreme-valued biomarkers (on average participants without depression had 1.96 [1.90, 2.01] extreme values, participants with depression had 2.25 [2.08, 2.43]; average difference between groups in number of extreme values = 0.248 [0.056, 0.440], Figure 2B-C).

**Figure 2:**
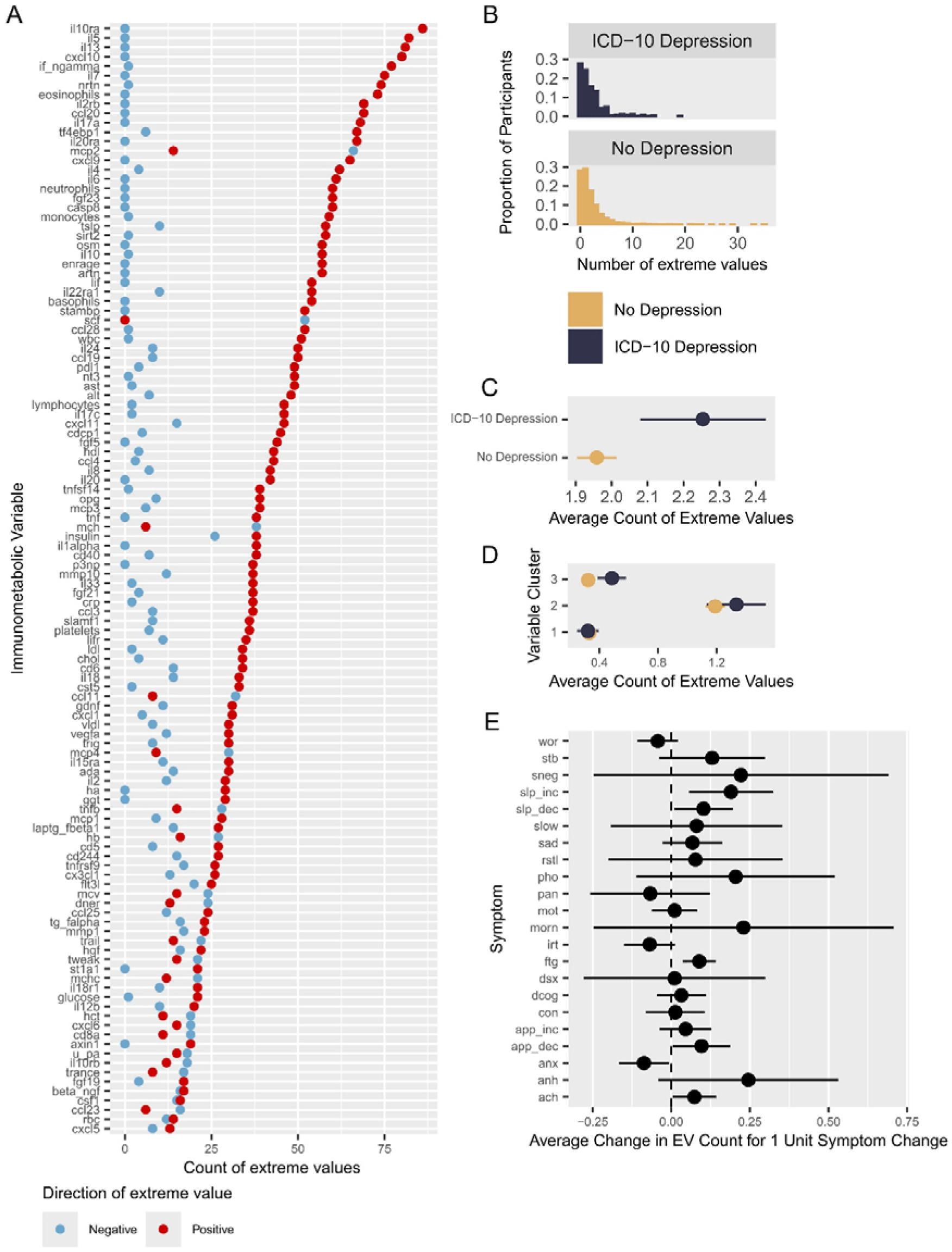
Extreme Value Distributions *A.* Count of total number of positive (red) and negative (blue) valued immuno-metabolic variables for each immuno-metabolic variables (refer to **Supplementary Table 1** for full immuno-metabolic variable definition) *B.* Histogram of proportion of participants with different counts of extreme values, split by ICD-10 depression diagnosis. *C.* Model-predicted average counts of extreme values per participant, split by ICD-10 depression diagnosis, with 95% confidence interval *D.* Model-predicted mean number of extreme values across immuno-metabolic variables, grouped by clusters identified in Figure 1, with 95% confidence intervals. *E.* Model predicted association between symptom score and extreme value count: average change in extreme value count associated with a 1 unit increase in the focal symptom score, with 95% confidence intervals. Symptoms as defined in Figure 1.

Next, to test if extreme variability in biomarker level in depression is specific to any biomarker cluster, we fit a model including immuno-metabolic cluster identity. We found an interaction between depression and cluster on the number of extreme values (χ^2^ =11.262, P=0.003). The number of extreme-valued biomarkers was higher in people with depression for biomarkers belonging to the positive association cluster (cluster 3; AME=0.104 [0.033, 0.174]), but not the other two clusters (AME for the negative association cluster 1 = −0.015 [−0.070, 0.040], and AME for low association cluster 2 = 0.067 [−0.085, 0.218]) (Figure 2D).

Finally, we fit models using scores on our symptom scales as predictors of number of extreme variables (Figure 2E**, Supplementary Table 2**). After correction for multiple comparisons, higher scores on the fatigue symptom scale (ftg) were associated with higher extreme immuno-metabolic variables: a 1 unit increase in ftg was associated with an average 0.089 (0.036, 0.142) increase in extreme values, P_FDR_ = 0.021.

### 3.5 Latent Components Underlying Symptom - Immuno-metabolic Biomarker Associations

Our clustering analysis identified that some immuno-metabolic variables had similar profiles of association with depression and anxiety symptoms, and conversely some symptoms had similar profiles of association with immuno-metabolic variables. We explored corelations within immuno-metabolic variables. Immuno-metabolic variables were generally positively correlated (**Supplementary** Figure 4A and C), with the average correlation between variables being 0.114, SD = 0.1248. Principal components analysis PCA) indicated one large principal component (PC) explained 16.7% of variance in immuno-metabolic variables, while 8 PCs together explained 50% of variance.

Applying the same analysis to individual depressive symptoms (**Supplementary** Figure 4B and D), we found symptoms were also highly correlated (average correlation = 0.235, SD = 0.144). PCA indicated one large component explained 54% of variance.

We used Partial Least Squares (PLS) regression to ask if there were latent variables that capture covariation between immuno-metabolic biomarkers and depression symptoms. We fit a PLS model predicting 22 symptoms from the combination of 25 depression symptom-associated positive and negative association cluster biomarkers we identified using our clustering analysis and our 5 covariates (sex at birth, BMI at age 24, daily smoking, AUDIT-C score, and self-reported physical health conditions).

The best fitting PLS model identified 2 latent components explaining 22.1% of variance in immuno-metabolic variables and 48.5% of variance in the depression variables. Component 1 (labelled “*Somatic-Inflamed”*) loaded on multiple somatic and depressive symptoms (the top 4 variables by loading being fatigue, decreased sleep, decreased motivation, and aches & pains); on the immuno-metabolic side this component loaded most highly on higher neutrophil count, WBC count, IL-6 levels as well as female sex, daily smoking, and higher BMI. By contrast, Component 2 (labelled “*Anxiety-Non-inflamed”*) loaded on higher worry and anxiety, and lower Adenosine Deaminase, Monocyte Chemotactic Protein 1 (MCP1), IL-6 and CRP levels (Figure 3A-D).

**Figure 3:**
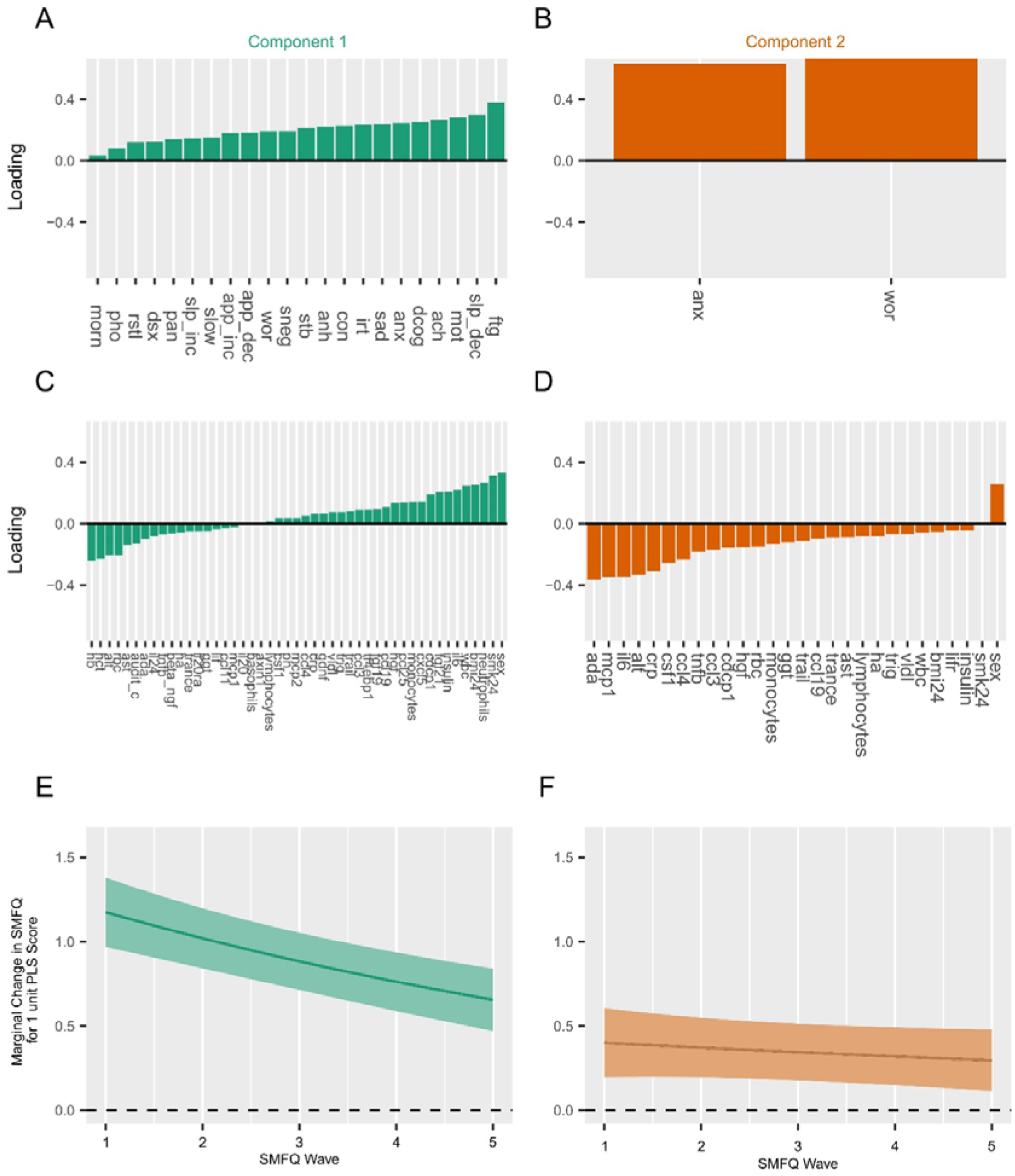
Latent components derived from Partial Least Squares analysis of immuno-metabolic variables and depressive symptoms A: PLS Component 1 Loadings on symptoms. B: As (A): Component 2 Loadings on symptoms C: PLS Component 1 Loadings on immuno-metabolic variables D: As (C): Component 2 Loadings on immuno-metabolic variables E: Plot of Marginal change in SMFQ total score associated with a 1 SD unit increase in PLS Component 1 score, with 95% confidence intervals, from generalised linear mixed model. F: As (F), marginal change in SMFQ total score for 1 unit increase in Component 2 score

Higher scores on both Components 1 and 2 were longitudinally associated with higher SMFQ depressive symptoms score at follow-up between ages 25 and 30 (Figure 3E-F). On average, a 1 SD unit increase in Component 1 score at age 24 was associated with a 1.170 [0.969, 1.380]) unit increase in SMFQ score at age 25, declining to a 0.656 [0.471, 0.841] unit increase around age 30. For component 2, a 1 SD unit increase in score at age 24 was associated with a 0.401 [0.195, 0.607] unit increase in SMFQ score, and a 0.298 [0.115, 0.480] unit increase around age 30.

### 3.6 Prediction of Depression and Specific Symptom Profiles using Immuno-metabolic Biomarkers

Finally, we tested how well the immuno-metabolic variables we identified as being associated with depression symptoms predict ICD-10 depression, using elastic net regression. Informed by our symptom-based analyses which suggested a distinction between somatic and mood symptoms vs anxiety symptoms, we also asked if immuno-metabolic variables were better at predicting specific symptom types within ICD-10 depression: depression *with* high levels of anxiety (high anxiety and worry; n=197/337) or depression *with* high somatic symptoms (fatigue, decreased sleep, low motivation and aches & pains; n=158/337), **Supplementary** Figure 5).

After nested cross-validation, models based on immuno-metabolic variables performed better than chance in predicting ICD-10 depression diagnosis (Balanced Accuracy=0.580; 95% CI = [0.568, 0.593]); Figure 4**, Supplementary Table 3.**

**Figure 4:**
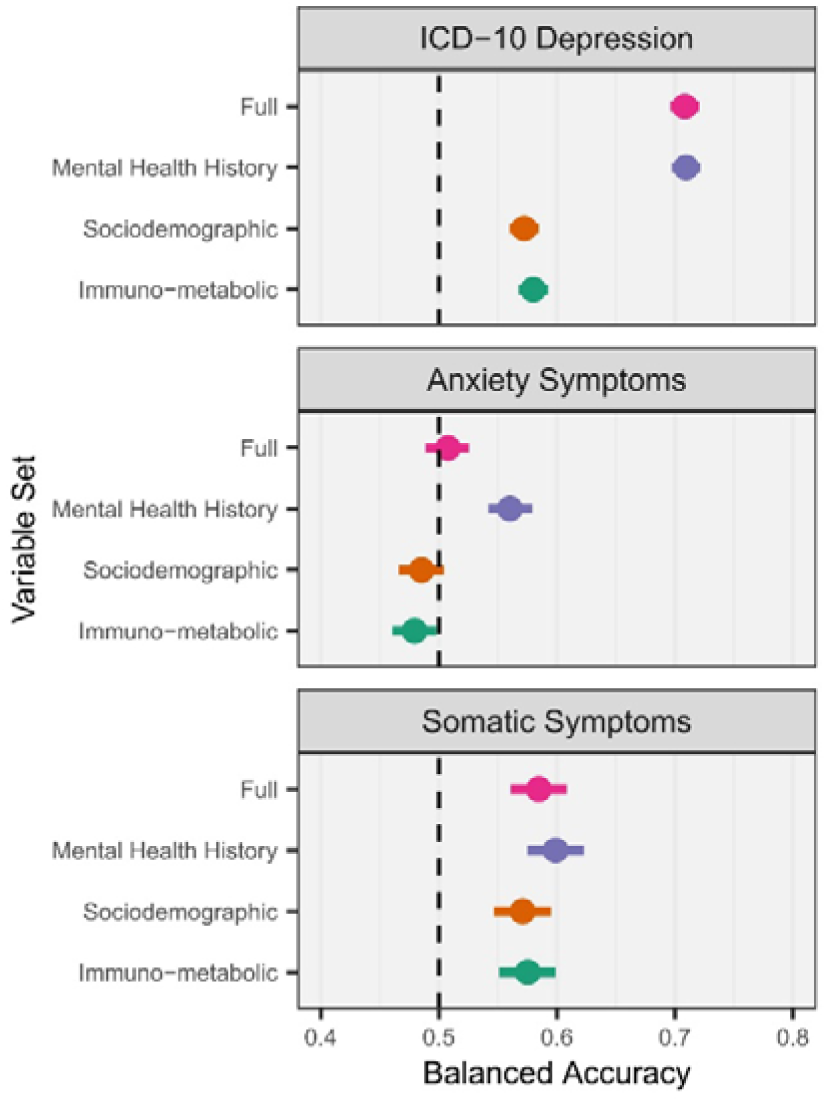
Performance of Machine Learning models predicting ICD-10 depression diagnosis and specific symptom profiles within individuals with ICD-10 depression Classification performance (balanced accuracy) of models trained with selected immuno-metabolic variables predicting the presence of ICD-10 depression, high anxiety symptoms within ICD-10 depression, or high somatic symptoms within ICD-10 depression, with 95% confidence intervals

Performance was similar for models trained using socio-demographic factors (0.572 [0.560, 0.585]), while a model trained with past mental health history (0.710 [0.697, 0.722]) and a model containing all variables (0.708 [0.696, 0.721]) performed better.

Within ICD-10 depression, immuno-metabolic biomarkers could be used to provide above-chance level predictions only of depression *with* high somatic symptoms (0.575 [0.551, 0.599]) whereas immuno-metabolic variables were not predictive of depression *with* high anxiety symptoms (0.479 [0.461, 0.498]). The difference in balanced accuracy for immuno-metabolic variables predicting high somatic and anxiety symptoms was 0.096 [0.066, 0.126]. By contrast, mental health history was predictive of higher levels of both anxiety and somatic symptoms within individuals with depression (**Supplementary Table 3)**.

## 4. Discussion

This study provides several insights into the associations between peripheral blood immune and metabolic biomarkers and depression symptoms. We found multiple biomarkers to be associated with a higher probability of depression in young adults as well as greater extreme variability in inflammatory biomarkers. The relationship between biomarkers and depression symptoms varied, with clusters of biomarkers showing distinct and opposing associations with somatic versus anxiety symptoms.

We identified two latent components potentially linking immuno-metabolic biomarkers and depression symptoms, one *somatic-inflammatory* component correlated with somatic symptoms, female sex, smoking and higher levels of systemic inflammatory measures, while the *anxious-non-inflamed* component related to anxiety, worry, female sex and lower levels of inflammatory biomarkers and growth factors. Higher somatic-inflamed component scores were associated with higher depressive symptoms severity in subsequent follow-up.

Machine learning models trained on immuno-metabolic biomarkers predicted ICD-10 depression diagnosis and depression with higher somatic symptoms, but not anxiety symptoms, better than chance. Predictive performance was similar to models trained on sociodemographic data, and to brain-imaging models (Winter et al., 2024). However, performance was at an absolute level considered poor (Alba et al., 2017).

Unlike most previous studies (Chu et al., 2019; Davyson et al., 2023; Milaneschi et al., 2021; Osimo et al., 2020), we examined a large number and variety of immuno-metabolic biomarkers (118 in total) including inflammatory proteins, cell count, lipids, and metabolites, which helped us identify key depression-related biomarkers (e.g., IL-6 and neutrophil count), reinforcing previous findings (Foley et al., 2023; Milaneschi et al., 2021; Osimo et al., 2020).

In addition, we leverage our larger set of immuno-metabolic variables to show that depression is associated more extreme valued biomarkers, as is the symptom fatigue, consistent with a system-wide disruption in immuno-metabolic homeostasis in young adults at age 24, and with a more somatic profile of symptoms. These differences were not related to current physical health conditions and occur well before the typical age of onset of cardiometabolic diseases, suggesting they are not purely a result of the presence of other conditions.

CRP has been associated with depression in meta-analysis (Osimo et al., 2020), but we did not reproduce this finding. There are several possible explanations for this apparent discrepancy. Mendelian randomisation studies have suggested a potential causal role of IL-6 in depression more consistently than CRP (Kappelmann et al., 2021; Perry et al., 2021b). CRP has also been found to have a small or protective effect against some mental disorders (Said et al 2022, Shi & Morrison 2025).

Longitudinal analysis of CRP has also suggested that there are specific trajectories of raised CRP across childhood which are predictive of mental disorders (Palmer et al., 2024). Therefore, although CRP is an archetypal clinical measure of systemic inflammation, single cross-sectional measurements may not be as strongly associated with depression as either upstream cytokines (e.g., IL-6), or longitudinal trends.

Some lymphocytes, including CD4^+^ helper T cells, CD19^+^ B Cells and NK Cells, have been found to be increased in depression (Foley et al., 2023). We did not find an association between lymphocyte count, or CD244, the NK cell receptor, and depression. However, CD244 is expressed on many immune cell types (Agresta et al., 2018). Our null finding could be due to limited resolution of measures used (WBC differential count) rather than flow-cytometry based subtyping.

We found several liver enzymes to be negatively associated with depression and anxiety symptoms. This is in contrast to a previous study which have found increased ALT in adults with higher depression scores (Zelber-Sagi et al., 2013). Our study population differs to the previous study, being substantially less male and much younger, meaning lower cumulative exposure to confounders such as alcohol. We also observed a positive correlation between liver enzymes, IL-6 and markers of red blood cell function. Lower Hb could be associated with poor health through multiple mechanism including chronic disease, poor dietary iron intake, blood loss and deficiencies in multiple vitamins (e.g. B12 and folate); therefore, lower liver enzymes could be a correlate of these markers of poor health and lower Hb, which also appeared to be associated with anxiety symptoms.

We also identified several novel potential biomarkers associated with depressive symptoms including CCL25, FGF19 and FGF21. Expressed in the gut, CCL25 has been associated with bipolar disorder (Göteson et al., 2022; Poletti et al., 2021), post-traumatic stress disorder (Zhang et al., 2020), bowel inflammation (Wurbel et al., 2011) and impaired intestinal barrier function (Jensen et al., 2023) in schizophrenia. FGF19 and FGF21 are linked to glucose and glycogen metabolism (BonDurant and Potthoff, 2018; Kir et al., 2011); FGF21 is associated with the risk of type-two diabetes (Post et al., 2023).

Taken together, our findings suggest that there may be distinct immuno-metabolic biotypes linked to specific symptom patterns in depression (somatic *vs* anxiety symptoms). These putative biotypes could potentially map on to other hypothecated depression biotypes, such as those based on brain imaging (Tozzi et al., 2024).

By assaying immuno-metabolic biomarkers, in combination with clinical history (Arribas et al., 2025; Zhang et al., 2024), brain imaging (Fu et al., 2024) or actigraphy (Price et al., 2025), patients could be stratified to give better information on prognosis or targeting specific treatments. For example, an important question for future research is whether patients with a somatic-inflammatory biotype could improve more with anti-inflammatory drugs compared to patients with an anxious-non-inflamed biotype.

Given the association we identified between somatic symptoms and FGF19 and FGF21, and these hormones and the effects of GLP-1 receptor agonists (Shao and Jin, 2022), which have recently been associated with numerous potentially beneficial mental health outcomes (Chen et al., 2024; Xie et al., 2025), another avenue might be to explore if depression with somatic symptoms is improved by GLP-1 receptor agonists.

Our study has several limitations. The Olink assay is widely used across proteomic studies in other cohort studies including UK Biobank (Sun et al., 2023), but gives results on an arbitrary scale. For IL-6, a high correlation between a clinical chemistry assay and Olink levels has been demonstrated in ALSPAC (Goulding et al., 2022). However, Olink may differ in target epitopes to other assays (Eldjarn et al., 2023), leading to only modest correlations with the SomaScan assay.

While the absolute values we report may not be generalisable, the pattern of change in relative levels of immuno-metabolic variables may be relevant to studies using other methods.

The ALSPAC study is not diverse in terms of ethnicity. Individuals who have continued to participate into early adulthood tend to be women, come from more educated and affluent backgrounds (Donnelly et al., 2022), and have a lower prevalence of depressive symptoms (Office for National Statistics, 2021) than the general UK population. Therefore, the generalisability of our findings to other populations may be limited and our conclusions need to be externally validated in populations with different characteristics, e.g. age, ethnicity and co-morbid health conditions. Larger population sizes are also necessary to improve and validate machine learning models.

Immuno-metabolic biomarkers were assayed at age 24. It is possible that longitudinal trends in biomarker levels derived from repeated measurement could yield more accurate prediction, but such datasets remain scarce e.g. (Palmer et al., 2024; Perry et al., 2021a).

Our longitudinal follow-up data was limited to SMFQ scores. The SMFQ does not include somatic symptoms. However, at present follow-up CIS-R data is not available from ALSPAC. Additionally, although our sample size is reasonably large, in a group of young adults a small number would be expected to develop psychiatric disorders other than depression, including bipolar affective disorder and psychotic disorders, but may be included in the depression group in the present data.

Future studies of longitudinal associations when more detailed psychiatric symptom data are available will be invaluable in evaluating the association of immuno-metabolic biomarkers with long-term trends in psychiatric symptoms.

Finally, our cross-sectional study was not designed, or able, to prove causal relationships or suggest mechanisms by which immune or metabolic dysfunction might impact the brain systems involved in the manifestation of the subjective experience of depression. Other study designs are necessary to explore this.

## 5. Conclusions

We found depression is associated with changes in multiple immuno-metabolic markers. Specific patterns of immuno-metabolic biomarkers are associated with differing subsets of depression symptoms (somatic-inflamed vs anxiety-non-inflamed), which are potentially relevant for future symptom severity. Immuno-metabolic biomarkers predict depression and high levels of somatic symptoms, but not anxiety symptoms, better than chance. Our results highlight the heterogeneity of depression and opportunities for immuno-metabolic biomarker-based subtyping, prediction and targeted intervention in future.

## Data Availability

Data from the ALSPAC study is available via a managed open access system: in line with ALSPAC data access policy, all raw data used for this work can be accessed by other investigators by making a request to the ALSPAC study executive (see the study website: http://www.bristol.ac.uk/alspac/researchers/access/). Available variables can be browsed via the ALSPAC data dictionary (http://www.bristol.ac.uk/alspac/researchers/our-data/) and variable search (https://variables.alspac.bris.ac.uk/). Analysis scripts are available online: https://github.com/NADonnelly/dep_infl

## Acknowledgements

We are extremely grateful to all the families who took part in the ALSPAC cohort study, the midwives for their help in recruiting them, and the whole ALSPAC team, which includes interviewers, computer and laboratory technicians, clerical workers, research scientists, volunteers, managers, receptionists and nurses. The views expressed are those of the authors and not necessarily those of the NIHR or the Department of Health and Social Care.

## Preprint

An early version of this manuscript was deposited as a preprint to medRxiv: https://doi.org/10.1101/2024.06.03.24308351

## CRediT authorship statement

Donnelly N: Conceptualisation; Study design; Software; Data Curation; Formal Analysis; Visualization; Writing – Original Draft; Writing – Reviewing & Editing Tsang RSM: Writing – Reviewing & Editing; Resources Foley E: Writing – Reviewing & Editing; Resources Fraser H: Writing – Reviewing & Editing; Resources Hanson A: Writing – Reviewing & Editing; Resources Khandaker GM: Conceptualisation; Study design; Writing – Reviewing & Editing; Funding acquisition; Supervision

