## Supplement for "Blood Immuno-metabolic Biomarker Signatures of Depression and Affective Symptoms in Young Adults"

### Supplementary Figures

#### Supplementary Figure 1

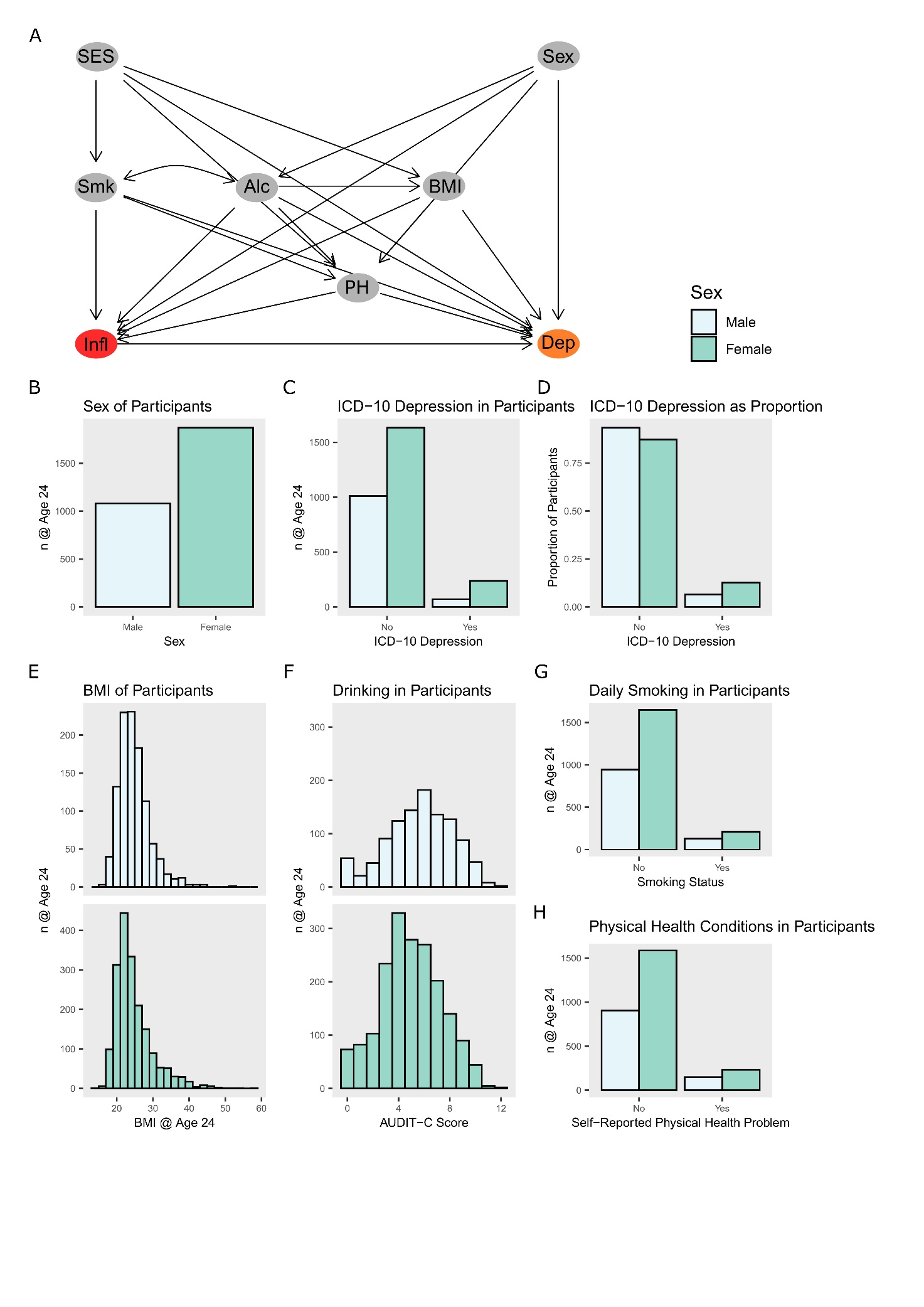

**Supplementary Figure 1: Dataset Overview for depression and key covariates**

A. *Study Directed Acyclic Graph: arrows (edges) illustrate proposed causal relationships. Inflammation (Infl, coloured red) is the proposed exposure; Depression (Dep, coloured orange) is the proposed outcome. Covariates, coloured grey, are: Socio-economic status (SES), Smoking (Smk), Alcohol use (Alc), Body Mass Index (BMI), Sex at birth (Sex) and Physical Health conditions (PH).*
B. *Count of participants at age 24, by sex at birth*
C. *Count of participants at age 24 by ICD-10 Depression Diagnosis and sex at birth*
D. *Proportion of participants by sex at birth, split by ICD-10 Depression Diagnosis*
E. *Distribution of BMI at age 24, by sex at birth*
F. *Distribution of Audit C scores at age 24, by sex at birth*
G. *Count of participants at age 24 by daily smoking status, split by sex at birth*
H. *Count of participants at age 24 who self-reported physical health problems*

#### Supplementary Figure 2

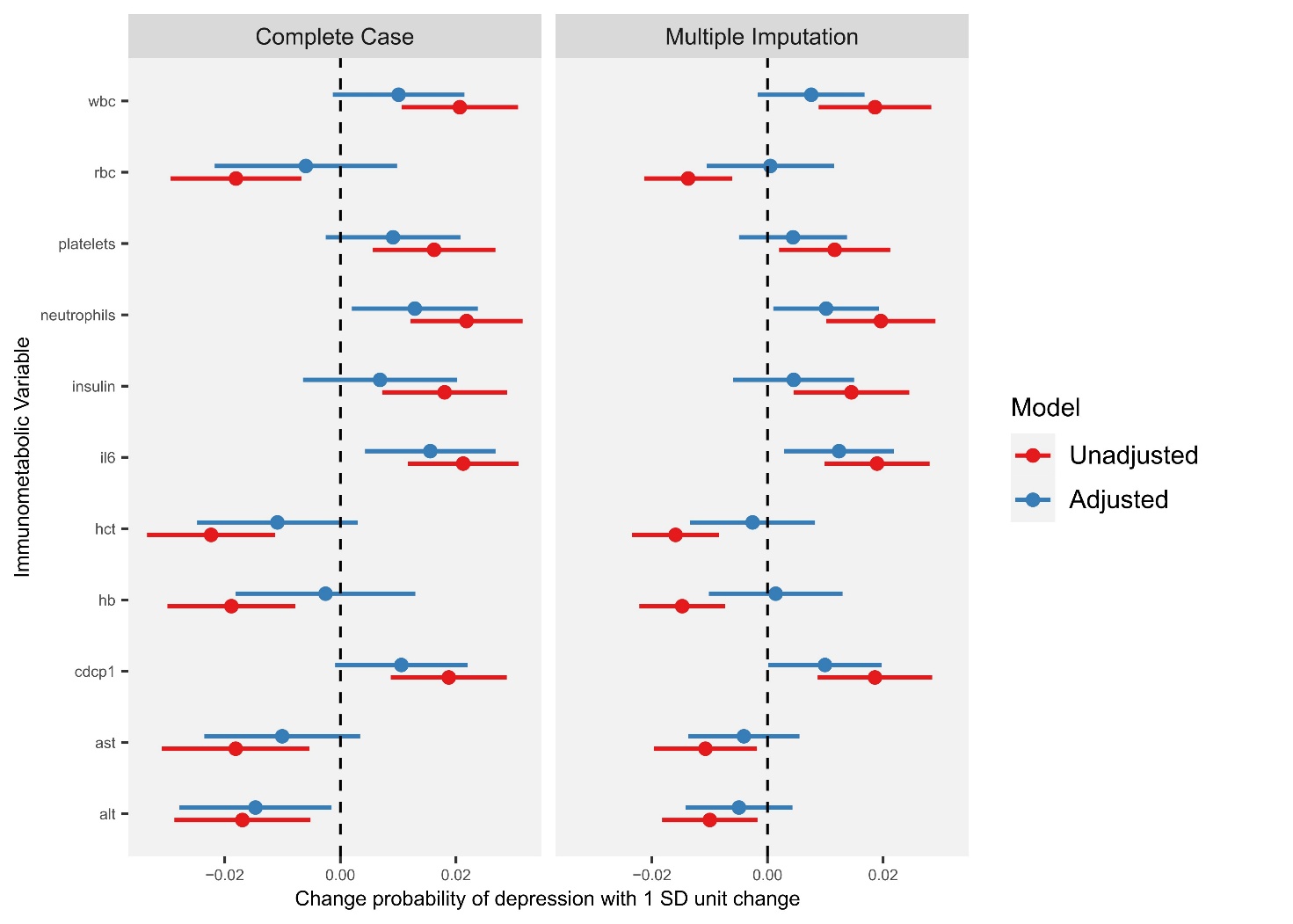

**Supplementary** **Figure 2: Association between immuno-metabolic biomarkers and depression at age 24 years in the ALSPAC birth cohort**

*This figure shows results from the analysis of complete case set (Left) and dataset after multiple imputation (Right). Results represent the average marginal effect and 95% confidence interval of a 1 SD unit change in the immuno-metabolic variable of interest and the probability of ICD-10 depression. Model 1 (red) is unadjusted for any covariates; Model 2 (blue) is adjusted for alcohol use, body mass index (BMI), self-reported physical health conditions, sex at birth and smoking status. Variable names: alt = Alanine aminotransferase; ast = Aspartate aminotransferance; cdcp1 = CUB domain-containing protein 1; hb = Haemoglobin; hct = haematocrit; il6 = Interleukin 6; insulin = random insulin; neutrophil = neutrophil count; platelets = platelet count; rbc = red blood cell count; wbc = total white blood cell count. See* ***Supplementary Table 1*** *for full variable descriptions.*

#### Supplementary Figure 3

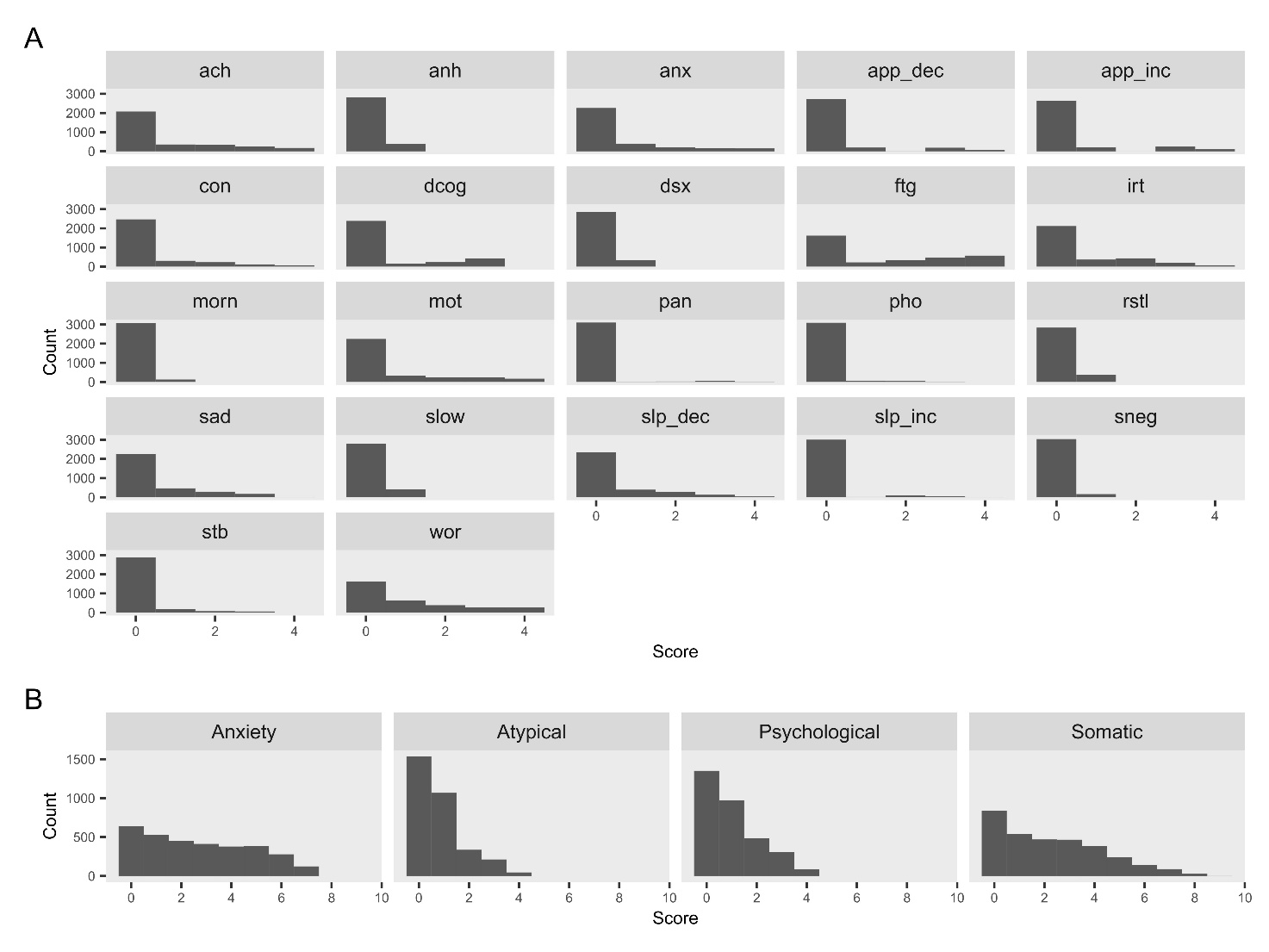

**Supplementary Figure 3: Distribution of symptom and domain scores**

*A: Histograms of distribution of scores for all participants on all 22 derived symptom scores: :* Aches & Pains (ach), Anhedonia (anh), Anxiety (anx), Appetite Decreased (app_dec), Appetite Increased (app_inc), Concentration Impaired (Con), Depressive Cognitions – feelings of guilt, inadequacy and helplessness (dcog), Decreased interested in sex (dsx), Fatigue (ftg), Irritability (irt), Morning low mood (morn), Motivation impairment (mot), Panic (pan), Phobias/specific fears (pho), Restlessness (rstl), Sadness/low mood (sad), Slowed movements (slow), Sleep Decreased (slp_dec), Sleep Increased (slp_inc), Self-neglect (sneg), Suicidal Thoughts (stb) and Worry (wor) *(see* ***Supplementary Methods*** *for derivations)*

*B: Histograms of distribution of scores of all participants for four symptom domain scales. Full derivation is given in the* ***Supplementary Methods***

#### Supplementary Figure 4

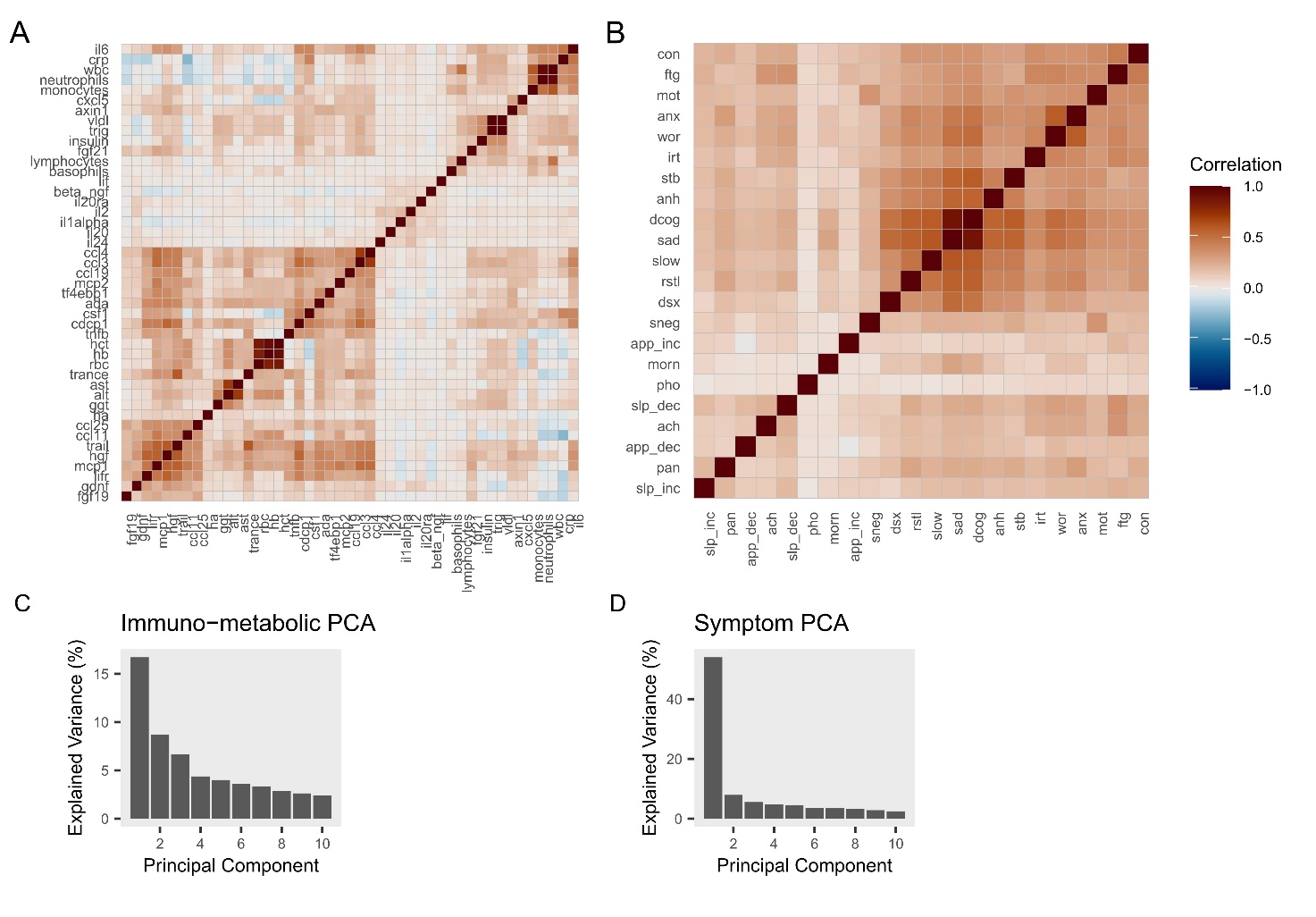

**Supplementary Figure 4: Variable correlations for immuno-metabolic variables and depression symptoms**

A: Correlation coefficients between all cluster 1 and 3 immuno-metabolic variables (red = higher correlation, blue = lower correlation). Note variables are sorted after application of a clustering method (“complete” method in R *hclust* function)

B: Correlation coefficients between all individual depression symptoms (see Supplementary Figure 3 for all symptom descriptions)

C: Histogram of component explained variables after applying Principal Components Analysis

#### Supplementary Figure 5

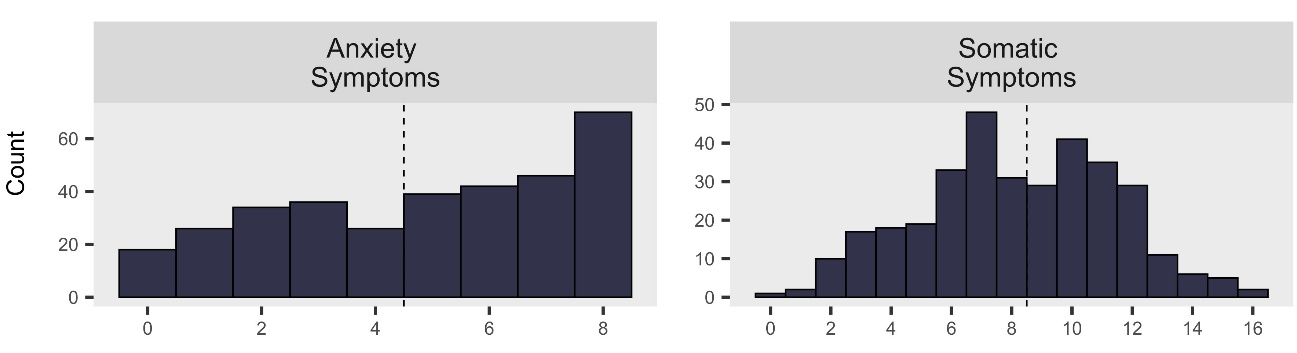

**Supplementary Figure 5: Distribution of anxiety and somatic symptom scores within participants with depression**

Histograms of all individuals with ICD-10 Depression. Dashed vertical lines indicate cut offs used to generate target variables for classification models.

### Supplementary Tables

#### Supplementary Table 1

**Supplementary Table 1**: All Immuno-metabolic Variables, mean values by sex (standard deviation) and depression associations (standardised mean difference and 95% confidence interval) are found in *supplementary_table_1.xlsx*

#### Supplementary Table 2

| Symptom | Abbreviation | Estimated change in extreme values for 1 unit change in symptom, (95% CI), P_FDR_ |
| --- | --- | --- |
| Aches & Pains | ach | 0.074 (0.005, 0.142), P = 0.14 |
| Anhedonia | anh | 0.245 (-0.042, 0.531), P = 0.266 |
| Anxiety | anx | -0.087 (-0.167, -0.007), P = 0.14 |
| Appetite Decreased | app_dec | 0.096 (0.005, 0.187), P = 0.14 |
| Appetite Increased | app_inc | 0.045 (-0.037, 0.128), P = 0.477 |
| Concentration Impaired | con | 0.013 (-0.081, 0.106), P = 0.825 |
| Depressive Cognitions | dcog | 0.032 (-0.046, 0.111), P = 0.575 |
| Decreased interested in sex | dsx | 0.011 (-0.278, 0.299), P = 0.943 |
| Fatigue | ftg | 0.089 (0.036, 0.142), P = 0.021 |
| Irritability | irt | -0.069 (-0.15, 0.012), P = 0.266 |
| Morning low mood | morn | 0.23 (-0.247, 0.707), P = 0.519 |
| Motivation impairment | mot | 0.01 (-0.062, 0.083), P = 0.825 |
| Panic | pan | -0.067 (-0.258, 0.124), P = 0.634 |
| Phobias | pho | 0.204 (-0.111, 0.52), P = 0.374 |
| Restlessness | rstl | 0.077 (-0.2, 0.354), P = 0.678 |
| Sadness | sad | 0.068 (-0.028, 0.163), P = 0.361 |
| Slowed movements | slow | 0.08 (-0.192, 0.353), P = 0.678 |
| Sleep Decreased | slp_dec | 0.103 (0.01, 0.196), P = 0.14 |
| Sleep Increased | slp_inc | 0.19 (0.056, 0.324), P = 0.06 |
| Self-neglect | sneg | 0.222 (-0.247, 0.691), P = 0.519 |
| Suicidal Thoughts | stb | 0.13 (-0.038, 0.298), P = 0.317 |
| Worry | wor | -0.043 (-0.107, 0.021), P = 0.374 |

#### Supplementary Table 3

**Supplementary Table 3:** Machine learning model performance after nested cross-validation

| Diagnosis/Symptom Set | Variable Set | Balanced Accuracy | AUROC | MCC |
| --- | --- | --- | --- | --- |
| ICD-10 Depression | Immuno-metabolic | 0.580 (0.568, 0.593) | 0.612 (0.6, 0.625) | 0.102 (0.084, 0.118) |
|  | Sociodemographic | 0.572 (0.560, 0.585) | 0.603 (0.59, 0.615) | 0.092 (0.075, 0.109) |
|  | Mental Health History | 0.710 (0.697, 0.722) | 0.781 (0.768, 0.794) | 0.277 (0.26, 0.295) |
|  | Full | 0.708 (0.696, 0.721) | 0.783 (0.77, 0.796) | 0.280 (0.263, 0.297) |
| Depression with high Anxiety Symptoms | Immuno-metabolic | 0.479 (0.461, 0.498) | 0.473 (0.453, 0.493) | -0.093 (-0.157, -0.031) |
|  | Sociodemographic | 0.485 (0.466, 0.504) | 0.481 (0.46, 0.502) | -0.081 (-0.149, -0.015) |
|  | Mental Health History | 0.560 (0.542, 0.579) | 0.592 (0.572, 0.612) | 0.125 (0.083, 0.168) |
|  | Full | 0.508 (0.489, 0.525) | 0.51 (0.49, 0.531) | 0.02 (-0.023, 0.065) |
| Depression with high Somatic Symptoms | Immuno-metabolic | 0.575 (0.551, 0.599) | 0.598 (0.571, 0.624) | 0.151 (0.103, 0.198) |
|  | Sociodemographic | 0.571 (0.547, 0.595) | 0.606 (0.58, 0.633) | 0.143 (0.095, 0.191) |
|  | Mental Health History | 0.599 (0.575, 0.623) | 0.653 (0.625, 0.678) | 0.198 (0.151, 0.247) |
|  | Full | 0.585 (0.561, 0.608) | 0.615 (0.588, 0.641) | 0.169 (0.124, 0.218) |

### Supplementary Methods

#### Clinical Datasets

##### Clinical Outcomes

Our primary outcome was International Classification of Diseases 10th Revision (ICD-10) Depression diagnosis, which was represented as a binary variable (non-depressed/depressed at the time of interview, combining all individuals with mild, moderate, and severe depression into the depressed group; ALSPAC variable **FKDQ1000**).

##### Symptom Scores

The following individual domain scores were created from the elements of the CIS-R Clinical Interview included in the ALSPAC Focus@24 Clinic (note YP = Young Person):

- Aches and Pains (0-4 scale) = sum of:
  - **FKDQ2620** (In past 7 days, number of days YP noticed this pain/ache) = “Four days or more” OR **FKDQ2720** (In past 7 days, number of days YP noticed this discomfort) = “Four days or more” [score 0/1]
  - **FKDQ2630** (In past 7 days, pain/ache lasted for >3 hours on a given day) = “Yes” OR **FKDQ2730** (In past 7 days, discomfort lasted for >3 hours on a given day) = “Yes” [score 0/1]
  - **FKDQ2640** (In past 7 days, the pain has been unpleasant) = “Unpleasant” OR “Very unpleasant” OR **FKDQ2740** (In past 7 days, the discomfort has been unpleasant) = “Unpleasant” OR “Very unpleasant” [score 0/1]
  - **FKDQ2650** (In past 7 days, pain has bothered YP when doing something interesting) = “Yes” OR **FKDQ2750** (In past 7 days, discomfort bothered YP when doing something interesting) = “Yes” [score 0/1]
- Anhedonia (0-1 scale) = sum of:
  - **FKDQ5030** (In past 7 days, YP has been able to enjoy things as much as usual) = “No, less enjoyment than usual” = 1; “No, did not enjoy anything” = 1 [score 0/1]
- Anxiety (0-4 scale) = sum of:
  - **FKDQ6300** (In past 7 days, number of days YP felt generally anxious/nervous) = “Four days or more” [score 0/1]
  - **FKDQ6310** (In past 7 days, how unpleasant anxiety/nervousness has been) = “Unpleasant” OR “Very unpleasant” [score 0/1]
  - **FKDQ6320** (In past 7 days, when anxious YP's heart raced/pounded) = “Yes” OR **FKDQ6330** (In past 7 days, when anxious YP felt dizzy) = “Yes” OR **FKDQ6340** (In past 7 days, when anxious YP felt nauseous/had a stomach ache) = “Yes” OR **FKDQ6350** (In past 7 days, when anxious YP had sweaty/shaky hands) = “Yes” OR **FKDQ6360** (In past 7 days, when anxious YP had difficulty catching breath) = “Yes” OR **FKDQ6370** (In past 7 days, when anxious YP had dry mouth) = “Yes” OR **FKDQ6380** (In past 7 days, when anxious YP had chest pain) = “Yes” OR **FKDQ6390** (In past 7 days, when anxious YP had numb/tingly hands or feet) = “Yes” [score 0/1]
  - **FKDQ6400** (In past 7 days, YP has felt anxious/nervous for >3 hours in a day) = “Yes” [score 0/1]
- Appetite Decreased (0-4 scale) = sum of:
  - **FKDQ2000** (past month, YP has noticed a marked loss in appetite) = “Yes” [score 0/1]
  - **FKDQ2010** (In past month, YP has lost any weight) = “Yes” [score 0/1]
  - **FKDQ2030** (Amount of weight lost In past month) = “Less than half a stone” = 1; “Half a stone or more” = 2 [score 0/1/2]
- Appetite Increased (0-4 scale) = sum of:
  - **FKDQ2100** (In past month, YP has noticed a marked increase in appetite) = “Yes” [score 0/1]
  - **FKDQ2110** (In past month, YP has gained any weight (if male)) = “Yes” OR **FKDQ2120** (In past month, YP has gained any weight (if female)) = “Yes” [score 0/1]
  - **FKDQ2140** (Amount of weight gained In past month) = “Less than half a stone” = 1; “Half a stone or more” = 2 [score 0/1/2]
- Concentration and Forgetfulness (0-4 scale) = sum of:
  - **FKDQ3520** (In past 7 days, number of days YP noticed concentration/memory problems) = “Four days or more” [score 0/1]
  - **FKDQ3530** (In past 7 days, YP could concentrate on TV show/newspaper/conversation) = “No” [score 0/1]
  - **FKDQ3540** (In past 7 days, concentration problems have stopped YP doing things) = “Yes” [score 0/1]
  - **FKDQ3560** (In past 7 days, YP has forgotten anything important) = “Yes” [score 0/1]
- Decreased interest in sex (0-1 scale) = sum of:
  - **FKDQ5110** (In past month, YP's interest in sex increased, decreased or stayed same) = “Decreased” [score 0/1]
- Depressive Cognitions (0-3 scale) = sum of:
  - **FKDQ5140** (In past 7 days, YP felt guilty/blamed themselves, even if not their fault) = “Sometimes” = 1; “Often” = 1 [score 0/1]
  - **FKDQ5150** (In past 7 days, YP felt they are not as good as other people) = “Yes, YP has not felt as good as others” = 1 [score 0/1]
  - **FKDQ5160** (In past 7 days, YP has felt helpless (e.g., about future)) = “Yes” = 1 [score 0/1]
- Early Morning Sadness (0-1 scale) = sum of:
  - **FKDQ5100** (In past 7 days, time of day felt more sad/depressed/uninterested) = “Worse in the morning” [score 0/1]
- Fatigue (0-4 scale) = sum of:
  - **FKDQ3020** (In past 7 days, number of days YP has felt tired) = “Four days or more” OR **FKDQ3120** (In past 7 days, number of days YP has been lacking in energy) = “Four days or more” [score 0/1]
  - **FKDQ3030** (In past 7 days, YP has felt tired for >3 hours on a given day) = “Yes” OR **FKDQ3130** (In past 7 days, YP felt lacking in energy for >3 hours on a given day) = “Yes” [score 0/1]
  - **FKDQ3040** (In past 7 days, YP has felt so tired they had to push self to do things) = “Yes, on one or more occasion” OR **FKDQ3140** (In past 7 days, YP felt so lacking in energy had to push self to do things) = “Yes, on one or more occasion” [score 0/1]
  - **FKDQ3050** (In past 7 days, YP has felt tired when doing things they enjoy) = “Yes” OR **FKDQ3150** (In past 7 days, YP has felt lacking in energy when doing things they enjoy) = “Yes” [score 0/1]
- Impaired Motivation (0-4 scale) = sum of:
  - **FKPE2020** (YP feels that they lack motivation when they have to do things) = “Yes, nearly always” = 1; “Yes, often” = 1 [score 0/1]
  - **FKPE2030** (YP feels that they spend all their days doing nothi) = “Yes, nearly always” = 1; “Yes, often” = 1 [score 0/1]
  - **FKPE2040** (YP feels that they are lacking in 'get up and go') = “Yes, nearly always” = 1; “Yes, often” = 1 [score 0/1]
  - **FKPE2090** (YP feels that they can never get things done) = “Yes, nearly always” = 1; “Yes, often” = 1 [score 0/1]
- Irritability (0-4 scale) = sum of:
  - **FKDQ4520** (In past 7 days, number of days YP felt irritable/short-tempered/angry) = “Four days or more” [score 0/1]
  - **FKDQ4530** (In past 7 days, YP felt irritable/short-tempered/angry for >1 hr in a day) = “Yes” [score 0/1]
  - **FKDQ4540** (In past 7 days, YP felt so irritable they wanted to shout at someone) = “Yes, but did not shout at someone” OR “Yes, and actually shouted at someone” [score 0/1]
  - **FKDQ4550** (In past 7 days, YP had arguments/rows/lost temper with someone) = “Yes” [score 0/1]
- Panic (0-4 scale) = sum of:
  - **FKDQ6710** (In past 7 days, frequency YP has felt panic): “Once” = 1; “More than once” = 2 [score 0/1/2]
  - **FKDQ6720** (In past 7 days, how unpleasant these feeling of panic have been) = “Unpleasant” OR “Unbearable, or very unpleasant” [score 0/1]
  - **FKDQ6730** (In past 7 days, the worst feeling of panic lasted >10 minutes) = “10 minutes or more” [score 0/1]
- Phobias (0-4 scale) = sum of:
  - **FKDQ6520** (In past 7 days, number of days YP felt anxious about most feared situation) = “Four or more days” [score 0/1]
  - **FKDQ6530** (In past 7 days, when anxious YP's heart raced/pounded) = “Yes” OR **FKDQ6540** (In past 7 days, when anxious YP felt dizzy) = “Yes” OR **FKDQ6550** (In past 7 days, when anxious YP felt nauseous/had a stomach ache) = “Yes” OR **FKDQ6560** (In past 7 days, when anxious YP had sweaty/shaky hands) = “Yes” OR **FKDQ6570** (In past 7 days, when anxious YP had difficulty catching breath) = “Yes” OR **FKDQ6580** (In past 7 days, when anxious YP had dry mouth) = “Yes” OR **FKDQ6590** (In past 7 days, when anxious YP had chest pain) = “Yes” OR **FKDQ6600** (In past 7 days, when anxious YP had numb/tingly hands or feet) = “Yes” [score 0/1]
  - **FKDQ6620** (In past 7 days, number of times YP avoided anxiety-inducing situations): “Between one and three times” = 1; “Four times or more" = 2 [score 0/1/2]
- Restlessness (0-1 scale) = sum of:
  - **FKDQ5120** (In past 7 days, when sad/uninterested YP has been restless/not sit still) = “Yes” [score 0/1]
- Sadness (0-4 scale) = sum of:
  - **FKDQ5010** (In past 7 days, YP felt sad/miserable/depressed) = “Yes” [score 0/1]
  - **FKDQ5040** (In past 7 days, no. of days YP felt sad/depressed/uninterested in things) = “Four days or more” = 1 [score 0/1]
  - **FKDQ5050** (In past 7 days, YP felt sad/depressed/uninterested for >3 hours in a day) = “Yes” [score 0/1]
  - **FKDQ5070** (In past 7 days, YP felt happier when something nice happened/in company) = “No, nothing cheered YP up” [score 0/1]
- Self-neglect (0-1 scale) = sum of:
  - **FKPE2080** (YP feels that they are neglecting their appearance/personal hygiene) = “Yes, nearly always” = 1; “Yes, often” = 1 [score 0/1]
- Sleep Decreased (0-4 scale) = sum of:
  - **FKDQ4010** (In past 7 nights, number of nights YP had sleep problems) = “Four nights or more” [score 0/1]
  - **FKDQ4030** (In past 7 days, number of nights spent >=3 hours getting to sleep) = “Four nights or more” [score 0/1]
  - **FKDQ4020** (past week, length of time getting to sleep on night with least sleep) = >1 hour to get to sleep [score 0/1]
  - **FKDQ4040** (In past 7 days, YP woke >2 hours early & couldn't get back to sleep) = “Yes” [score 0/1]
- Sleep Increased (0-4 scale) = sum of:
  - **FKDQ4010** (In past 7 nights, number of nights YP had sleep problems) = “Four nights or more” [score 0/1]
  - **FKDQ4130** (In past 7 days, number of nights YP slept for >3 hours more than usual) = “Four nights or more” [score 0/1]
  - **FKDQ4120** (In past 7 days, length of time over-slept on the night with most sleep) =“Between 15 minutes and 1 hour” = 1; “Between 1 and 3 hours " = 2; “Three hours or more” = 2 [score 0/1/2]
- Slowing (0-1 scale) = sum of:
  - **FKDQ5130** (In past 7 days, when sad/uninterested YP has done things slower than usual) = “Yes” [score 0/1]
- Suicidal Thoughts (0-3 scale) = sum of:
  - **FKDQ5170** (In past 7 days, YP has felt that life is not worth living) = “Sometimes” = 1; “Often” = 1 [score 0/1]
  - **FKDQ5180** (In past 7 days, YP has had thoughts of harming themselves) “Yes” = 1 [score 0/1]
  - **FKDQ5190** (In past 7 days, YP has thought about a way to kill themselves) “Yes” = 1 [score 0/1]
- Worry (0-4 scale) = sum of:
  - **FKDQ6020** (In past 7 days, number of days YP has worried about things) = “Four days or more” [score 0/1]
  - **FKDQ6030** (YP feels that that have been worrying too much) = “Yes” [score 0/1]
  - **FKDQ6040** (In past 7 days, how unpleasant worrying has been) = “Unpleasant” OR “Very unpleasant” [score 0/1]
  - **FKDQ6050** (In past 7 days, YP has worried for >3 hours in a day) = “Yes” [score 0/1]

##### Symptom Domains

The following symptom domains were derived from the ALSPAC CIS-R Interview, based on mapping CIS-R variables to their closest equivalent in (Milaneschi et al., 2021), which mapped symptoms between the PHQ-9 and GAD-7 to the Inventory of Depressive Symptomatology (IDS-SR30); see **Supplementary Table 1** in that publication (note YP = Young Person):

- Atypical Depressive Symptoms: sum of (1) **FKDQ2100** (In past month, YP has noticed a marked increase in appetite), (2) [**FKDQ2110** (In past month, YP has gained any weight (if male)) OR **FKDQ2120** (In past month, YP has gained any weight (if female))], (3) **FKDQ4100** (In past month, sleeping more than usual has been a problem) AND (4) **FKDQ3100** (In past month, YP feels that they have been lacking in energy), where each item was dichotomised into a single 0/1 score for presence/absence of the symptom. This gave a score on a 0 – 4 scale.
- Somatic Symptoms: sum of (1) **FKDQ4000** (In past month, YP had problems getting to sleep or back to sleep), (2) **FKDQ4100** (In past month, sleeping more than usual has been a problem), (3) **FKDQ3000** (In past month, YP has noticed that they have been getting tired), (4) **FKDQ3100** (In past month, YP feels that they have been lacking in energy), (5) **FKDQ2000** (In past month, YP has noticed a marked loss in appetite), (6) **FKDQ2100** (In past month, YP has noticed a marked increase in appetite), (7) **FKDQ3500** (In past month, YP has had problems concentrating), (8) **FKDQ5120** (In past 7 days, when sad/uninterested YP has been restless/not sit still) AND (9) **FKDQ5130** (In past 7 days, when sad/uninterested YP has done things slower than usual:), where each item was dichotomised into a single 0/1 score for presence/absence of the symptom. This gave a score on a 0 - 9 scale.
- Psychological Symptoms: sum of (1) **FKDQ5020** (In past month, YP has been able to enjoy things as much as usual [reverse coded]), (2) **FKDQ5000** (In past month, YP had a spell of feeling sad/miserable/depressed), (3) **FKDQ5150** (In past 7 days, YP felt they are not as good as other people) AND (4) **FKDQ5180** (In past 7 days, YP has had thoughts of harming themselves), where each item was dichotomised into a single 0/1 score for presence/absence of the symptom. This gave a score on a 0 - 4 scale.
- Anxiety Symptoms: sum of (1) **FKDQ6100** (In past month, YP has felt anxious/nervous), (2) FKDQ6000 (In past month, YP worried more than needed to about things), (3) FKDQ6030 (YP feels that that have been worrying too much), (4) **FKDQ6110** (In past month, YP's muscles felt tense or could not relax), (5) **FKDQ5120** (In past 7 days, when sad/uninterested YP has been restless/not sit still), (6) **FKDQ4500** (In past month, YP has felt irritable/short-tempered with others) AND (7) FKDQ6200 (In past month, YP felt anxious about situations where no real danger), where each item was dichotomised into a single 0/1 score for presence/absence of the symptom. This gave a score on a 0 - 7 scale.

#### Statistical Analysis

##### Directed Acyclic Graph and Covariate Selection

Prior to statistical analysis, we drew a Directed Acyclic Graph (DAG), explicating putative causal associations between our independent variable of interest (inflammation), dependent variable (depression or depressive symptoms) and relevant covariates **(Supplementary Figure 1A**). We drew out the DAG using Daggity (daggity.net) and used the *daggity* R package to identify the minimum adjustment set of covariates to adjust statistical models for (Tennant et al., 2021). This identified a set of key covariates for adjustment: sex at birth, BMI at age 24, smoking, alcohol use and physical health conditions.

##### Multiple Imputation

As a sensitivity analysis we used multiple imputation (MI) to impute missing data in our age 24 dataset. MI was performed on data from participants who either had blood sample data processed by Olink at age 9 or age 24 (total n = 4164). All age 24 blood variables and CIS-R-derived outcomes were imputed. We used age 9 blood variables, CIS-R mental health variables from age 18, covariates, and short mood and feeling questionnaire (SMFQ) results between ages 9 and 23 as auxiliary variables.

We used age 9 immuno-metabolic variables as auxiliary variables for age 24 variables as we found most of the age 9 blood measures had significant correlations with their age 24 counterparts (99/103 measures had significant [P_FDR_ < 0.05] correlations, the exceptions being tf4ebp1, stambp, axin1 and il1alpha), with a mean R^2^ value of 0.097, SD 0.091).

MI models were fit using the R *mice* package using 50 total imputations and 20 iterations, with random forest models as the imputation method. This imputation method fits random forest models to predict each variable using all other variables; random forest models have the advantage of being able to model both categorical and continuous variables and can capture both linear and non-linear relationships between predictors, providing theoretical advantage over other imputation methods (Waljee et al., 2013).

Models as described in the main methods were then fit to the full set of imputed data and model outputs were pooled using Rubin’s rules. The imputed dataset contained more males than the complete case data (43% compared to 37%), potentially addressing a source of bias in our complete case data.

##### Machine Learning Modelling

Elastic net regression was selected in preference to other machine learning models as it includes regularisation (a mixture of L1 and L2 i.e. LASSO and Ridge Regression) which allows for selection of only relevant variables. Additionally, as a form of logistic regression, model coefficients are readily interpretable as variable importances (compared to more complex black box machine learning methods); models can be fit quickly; and other more complex machine learning methods (e.g. random forests, neural networks or boosting) may require more data to give reliable estimates (i.e. hundreds of cases of depression per predictor variable included (van der Ploeg et al., 2014)), meaning that elastic net models were more likely to produce reliable and stable estimates in our dataset with only a few hundred cases of depression, but a wide pool of potential predictors.

Elastic net regression models hyperparameters (penalty and mixture) were selected by nested cross-validation (5-fold CV repeated 4 times to give 20 outer loops, inner training data were produced by 20 bootstraps from the inner training data). The search space for the penalty parameter was -10 to 0 on the log­_10_ scale; the search space for the mixture parameter was 0.05 – 1 (where 0 is a pure Ridge regression model and 1 is a pure LASSO regression model).

Within each outer cross-validation loop data were pre-processed as follows: an extreme value model for all cluster 3 immuno-metabolic variables was generated using the non-depressed participants in the outer loop training data, then this model was applied to depressed individuals in the outer loop training data, and all individuals in the outer loop testing data. Next, categorical variables (e.g. sex, daily smoking, maternal social class) were dummy coded, all continuous predictors were z-transformed and missing predictor data was imputed using K-Nearest Neighbours Imputation with five neighbours. Following these steps, to account for substantial class imbalances in outcomes (e.g. only ~10% of participants met criteria for ICD-10 depression), in model training only, synthetic oversampling of cases was used to produce balanced datasets using the ADASYN algorithm (He et al., 2008).

During training, in each outer loop, model classification performance on the held-out outer data in each fold was evaluated using balanced accuracy (*(sensitivity + specificity)/2) to* allow comparison with previous studies (e.g. (Winter et al., 2024)). We also calculated the area under the receiver-operator curve (AUROC) and Matthew’s Correlation Coefficient to facilitate comparison with other studies.
